# Electronic data capture for large scale typhoid surveillance, household contact tracing, and health utilisation survey: Strategic Typhoid Alliance across Africa and Asia

**DOI:** 10.1101/2020.03.02.20030056

**Authors:** Deus Thindwa, Yama G Farooq, Mila Shakya, Nirod Saha, Susan Tonks, Yaw Anokwa, Melita A Gordon, Carl Hartung, James E Meiring, Andrew J Pollard, Robert S Heyderman, on behalf of The Strategic Typhoid alliance across Africa and Asia consortium

## Abstract

**Background:** Electronic data capture systems (EDCs) have the potential to achieve efficiency and quality in collection of multisite data. We quantify volume, time, accuracy and costs of an EDC using large-scale census data from the STRATAA consortium, a comprehensive programme assessing population dynamics and epidemiology of typhoid fever in Malawi, Nepal and Bangladesh to inform vaccine and public health interventions.

**Results:** A census form was developed through a structured iterative process and implemented using Open Data Kit Collect running on Android-based tablets. Data were uploaded to Open Data Kit Aggregate, then auto-synced to MySQL-defined database nightly. Data were backed-up daily from 3 sites centrally, and auto-reported weekly. Pre-census materials’ costs were estimated. Demographics of 308,348 individuals from 80,851 households were recorded within average of 14.7 weeks range (13-16) using 65 fieldworkers. Overall, 21.7 errors (95% confidence interval: 21.4, 22.0) per 10,000 data points were found: 13.0 (95% confidence interval: 12.6, 13.5) and 24.5 (95% confidence interval: 24.1, 24.9) errors on numeric and text fields respectively. These values meet standard quality threshold of 50 errors per 10,000 data points. The EDC’s total variable cost was estimated at US$13,791.82 per site.

**Conclusions:** In conclusion, the EDC is robust, allowing for timely and high volume accurate data collection, and could be adopted in similar epidemiological settings.

## Background

Use of electronic data capture systems (EDCs) for health research has increased since Apple’s launch of the first handheld device in 1993 [1], and for observational studies and clinical trials is beginning to replace paper-based data collection methods. Paper-based systems have the advantage that they provide a hard copy source document but are characterised by high inaccuracies, substantial omissions, longer data turnaround time, longer data entry time, and high incremental costs both during the data collection and subsequent entry into an electronic database [2–6]. The advantages of EDC include built-in global positioning system (GPS) locator that automatically capture geographical coordinates thus minimizing transcription errors from external GPS locators; password-locked tablets and data encryption that maintain participant data confidentiality; required checks that prevent data omissions; range checks and data type checks that prevent typographical errors; skip patterns that provide logical responses; barcode technology that automates entry of unique identification; timestamps that provide a means to monitor work rate; and internet connectivity that ensures availability of real-time data [3, 4, 7, 8]. Despite these benefits [9], there is limited description of the performance of EDCs for large-scale or multisite surveys in low and middle-income countries.

Each year, an estimated 9.9-24.2 million typhoid fever cases occur from low and middle income countries resulting in approximately 75,000-208,000 deaths [10, 11]. However, although essential to build a public health case for disease control efforts such as vaccination and provision of clean water, sanitation and hygiene, obtaining reliable estimates for the burden of disease at national and sub-national level is difficult [12]. This requires collection of high quality field demographic, mapping, epidemiological, and clinical and laboratory data at scale from both hospital and community-based survey studies [13]. Interestingly, the collection of such quality data is hindered by complexities of dilapidated health facilities, overcrowding, unstructured housing or slums, and illiteracy [14].

We present an open source-based EDC, designed to overcome data quality complexities, and evaluate the efficiency, quality, and costs of the EDC by measuring volume, time, accuracy, and material costs using multisite census data collected from sub-Saharan Africa and Asia. The EDC was developed and implemented within the Strategic Typhoid alliance across Africa and Asia (STRATAA), a comprehensive programme which is assessing population dynamics and epidemiology of typhoid fever in Malawi, Bangladesh and Nepal to inform design of vaccine and public health interventions.

## Design, development and implementation

### Study setting and participants

The census component of the STRATAA study aimed to collect demographics from approximately 100,000 individuals, of all ages, in each of the three sites, to form the sampling frame for subsequent sub-studies. More details of the STRATAA study design and participants have previously been described [13]. In brief, the three sites, one in each country, were selected based on high known burden of enteric fever, differing epidemiological patterns and previous ability to deliver paper-based studies of high participant volume and logistical complexity.

### Electronic census report form and data cleaning procedure

An electronic census report form (eCRF), uniform to all sites, was developed through a structured iterative process. An eCRF comprised household- and individual-level questions. The eCRF data fields reflected a range of data types including integers to capture census team identifier, interviewer identifier, phone numbers of key respondent and older household members, household member number, and age; decimal to capture GPS points; alphanumeric to capture household unique identifier (barcode); texts to capture ward/traditional authority name, community/district name, physical address, respondent name, respondent relationship to head of household, respondent position in the household, head of household name, household member name, household member tribe/ethnicity, household member relationship to head, marital status, spouse name, education levels, employment status, mother’s name, and father’s name; characters to capture study site, household occupancy status, consent status, study information access status, sex, and school attendance status; and dates to capture household visit date and date of birth of each household member.

To ensure ultimate generation of error-free data, the eCRF data fields were designed with quality control tools such as dropdown menus, range checks, choice fields, skip patterns, required checks, double-data entry checks, systematic auto-numbering, preloading, and looping. However, due to other internal and external limitations of the EDC, we further built external database queries based on the Structured Query Language (SQL) to track potential data entry errors that might have arisen beyond EDC’s control. External SQL queries were aimed to expose persistent error sources which included duplication of study household identifiers (barcode); duplication of entire individual demographics; barcode decoding errors during scan; illogical ages or date of births of children relative to parents; incorrect household visit dates relative to tablet system date; misspellings of traditional authority names/ward numbers, physical addresses, respondent names, and household members names; missing GPS points; inaccurate GPS points relative to the household; and mismatches between community names and GPS points. After running the external SQL queries on the census database table and identifying the errors, each correction of an error by the data officer triggered an automatic log to an audit-trail table with entries (table’s column names) that included table name with error, action on an error (update, insertion, or deletion), individual/household barcode identifier with an error, field name with an error, old value, new value, timestamp, and a user’s name modifying an error. This generated a single row in an audit-trail table for each single error that was modified in the original census table. Errors corresponding to GPS points were specifically identified through sub-setting and importing GPS points (longitude, latitude, and altitude) from the census table into Google Earth Pro software (Google LLC, Mountain View, California, USA) as a keyhole markup language file, and then mapping the GPS points on the overlay of community boundaries’ and households’ satellite images. Once a GPS point was not mapped within 5 meters at 10% accuracy of the household or within the community boundary, it was considered a mapping error, and corrected through remapping in the field and updating it in the census table thereby triggering an audit-trail table error record. All the other errors exposed by the external SQL queries were investigated thoroughly in the field before corrections could be applied to the census table and subsequently auto-logged into the audit-trail table. The maximum number of visits to the household prior declaring the household vacant or errors permanently unresolved was twice. We provide the flow diagram of the eCRF in (Fig. 1), whereas the technical details of the extensible markup language code used to create an eCRF, and the SQL code used to create the audit-trail table and triggers to the audit-trail table have been publicly shared through GitHub (GitHub Inc, San Francisco, California, USA) [15].

**Figure 1.**
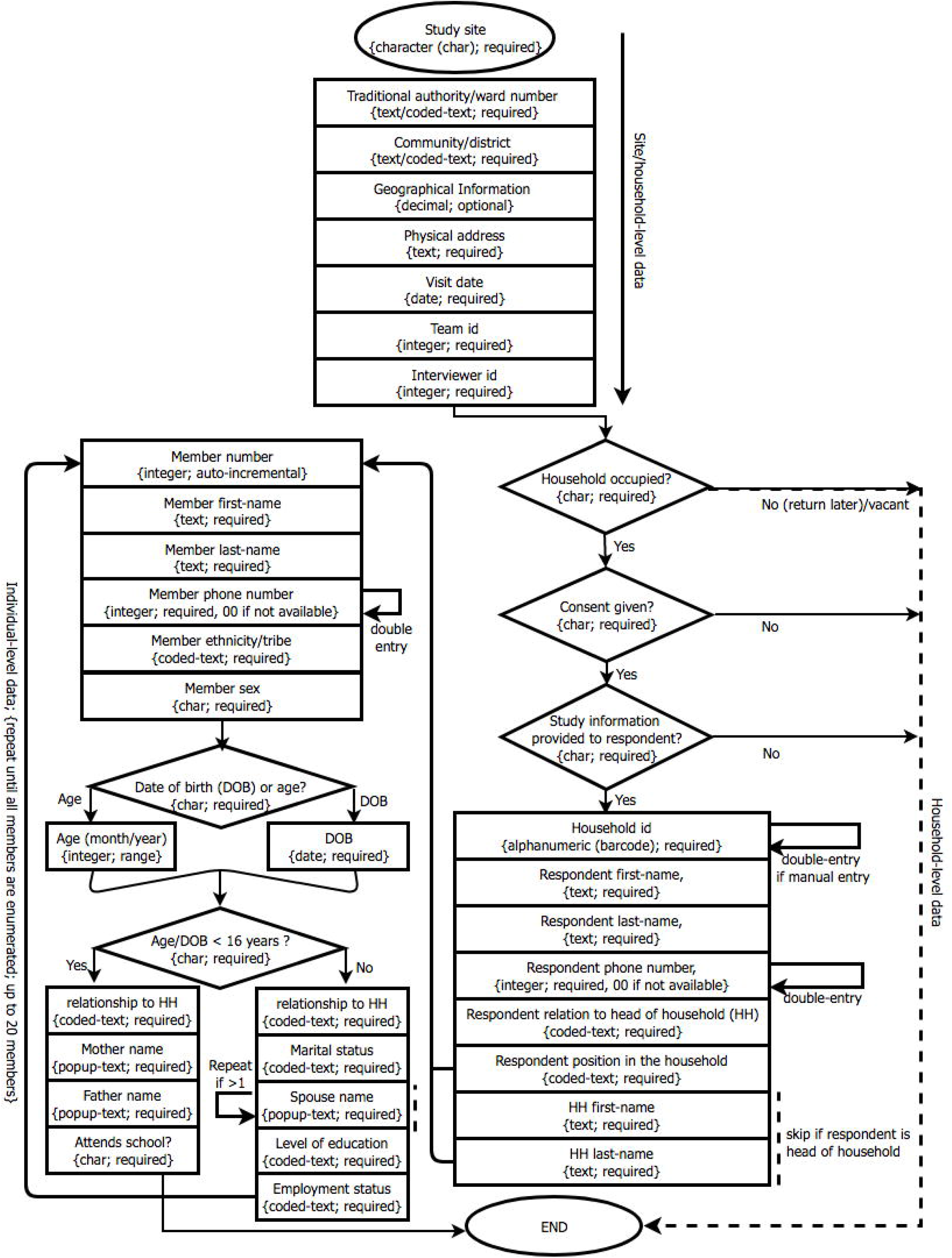
Electronic census report form (eCRF) flowchart.

### Electronic data capture system

We designed a uniform EDC using combined open-source tools; Open Data Kit (ODK) software (Nafundi, Seattle, Washington, USA) [16–18], and MySQL relational database management system (Oracle Corporation, Redwood city, California, USA) [19]. The eCRF was customized in ODK Collect and uploaded onto Android-based Asus ZenPad (AsusTek Computer Inc., Taipei, Taiwan), and Samsung (Samsung group, Seoul, South Korea) tablets. Then data were collected in the field during the day and temporarily saved in the tablet’s memory. At the end of each day, tablets were returned to the base STRATAA data office and data were uploaded from the tablet’s memory to ODK Aggregate server via a secure wireless network technology. Tablets were then charged overnight at the base data office in preparation for use on the next day. For every scheduled time of the night, data automatically synchronized from ODK Aggregate server to MySQL-defined database, set up for four main reasons; first, to facilitate corrections of inconsistencies beyond ODK validations (e.g. all persistent error sources mentioned above) and auto-audit the corrections; second, to ensure homogeneous database structure across sites in order to facilitate multisite dataset merging, and to preserve meaningful variables (excluding metadata generated by ODK software) in order to provide intuitive datasets to epidemiologists and statisticians; third, to generate automated reports using SQL; and last, to allow automated back-up of cleansed data from MySQL-defined database to external storage devices. The EDC also allows daily comma-separated value and anonymized data format to securely and automatically synchronize from each site’s ODK Aggregate server to a central repository. Conversely, the comma-separated value data format, from MySQL-defined database, are sporadically exported back to tablet’s ODK media folder to enable data preloading for sub-sequent sub-studies (Fig. 2). Technical details of the scripts for synchronizations, and creation of table structures and triggers have been publicly shared through GitHub [15].

**Figure 2.**
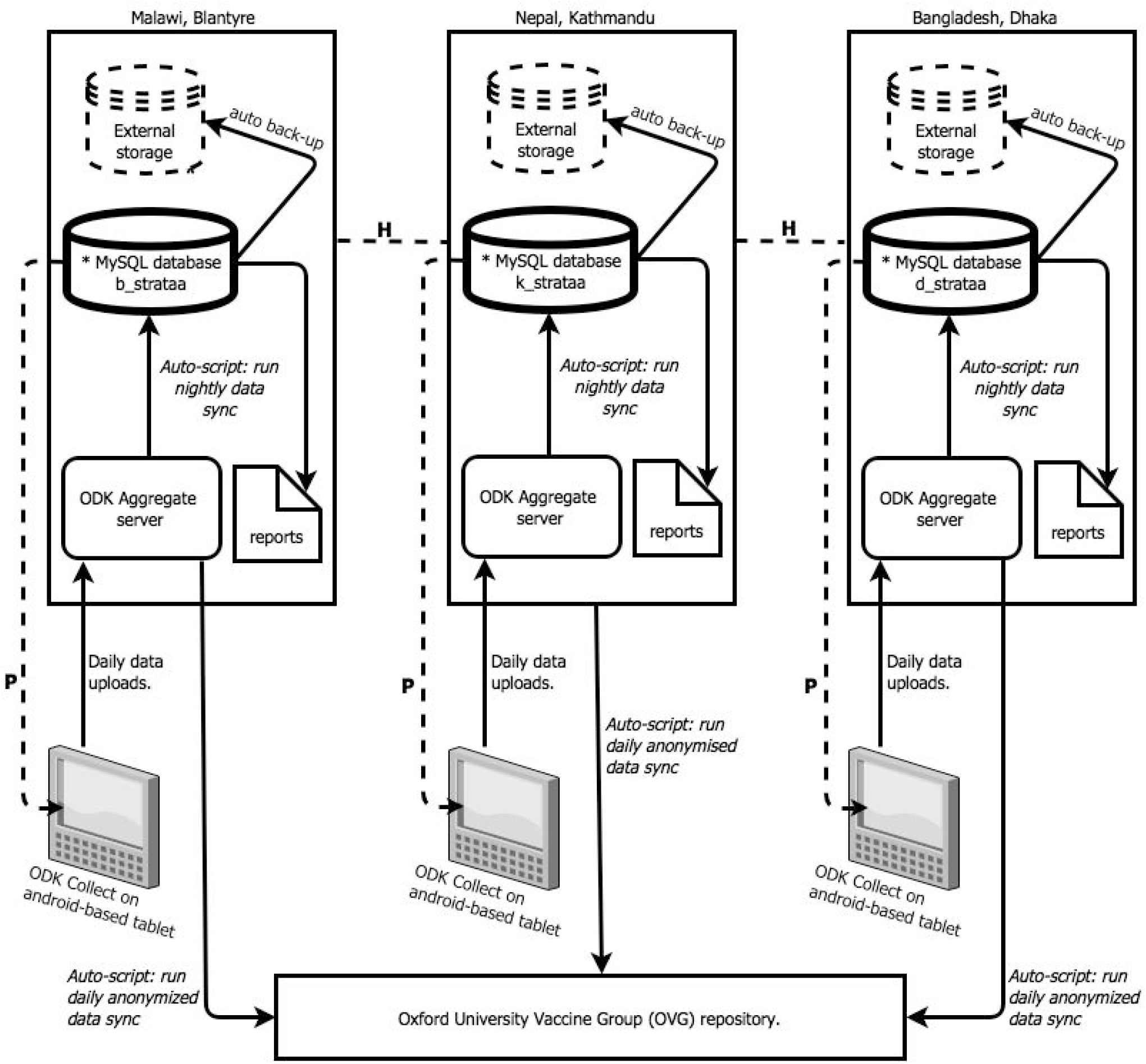
Electronic data capture system for a multisite study. MySQL-defined databases b_strataa, k_strataa, and d_strataa have homogeneous structures (*) e.g. table columns, data types, triggers or views. Data from MySQL-defined database table are exported back to Android-based tablet enabling data preloading for subsequent sub-studies (P). Homogeneous databases across sites merge enabling multisite data analyses (H).

### Pre-census time, costs, and training

We estimated time and costs required to attain the following census-related materials or complete census activities; tablets (including screen protectors, and protective covers), desktop server computers, network devices, barcodes, development of eCRF, training of field workers, replacement of broken tablets, and backpacks. We did not assess other operational costs because of uncertainty e.g. electric power to servers, charging tablets, and electronic data synchronization. We trained fieldworkers and assessed their suitability to conduct census by administering a practical mock test and then selecting best performers. Moreover, five weeks post-census implementation, we retrained fieldworkers based on calculated individual performances on data quality and data collection speed.

### Statistical analysis and visualization

We estimated the error rates, after running external SQL queries but prior to data cleaning, by dividing the total number of errors observed by the total number of data points (≈ all expected errors). A data point was defined as a discrete unit of information that could possibly be obtained from each member of the population after administering an eCRF e.g. If an eCRF had (*n*) number of unique questions, with each question corresponding to a variable (*X*_*i*_), for (*N*) number of respondents, then the total data points for eCRF would be 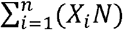. In our calculations, data points for household and individual-level variables were calculated separately and summed up. The reason was that household-level questions were answered by a key informant (head of household or respondent ≥ 18 years old) while individual-level questions were hypothetically answered by all household members (represented by a key informant). Exact binomial confidence intervals were used to estimate error rates. Data entry speed and accuracy by fieldworkers were combined into a single merit in order to measure their performance [20]. For each fieldworker, we standardized the data entry speeds (*z*_*s*_) and errors (*z*_*e*_), and assigned more weight to data entry speed (60%) than errors (40%) given the background that the EDC was robustly developed to prevent most data entry errors, thus, speed was more important. The final data entry speed-accuracy trade-off was calculated using the formula (*SAT* = −*z*_*s*_ ∗ 0.6 − *z*_*e*_ ∗ 0.4) where *z*_*s*_ (*s* − *μ*_*s*_)/*δ*_*s*_ and *z*_*e*_ = (*e* − *μ*_*e*_)/*δ*_*e*_(*s*) is the total speed for all data entries per field worker, (*δ*_*s*_) is the mean speed for all fieldworkers, (*e*) is the speed standard deviation, (*e*) is the total number of errors per field worker, (*μ*_*e*_) is the mean error for all fieldworkers, and (*δ*_*e*_) is the error standard deviation. In addition, we used Wilcoxon Signed-Rank Test for paired samples pre-versus post-retraining in order to measure any statistical difference in the number of errors committed, and determine whether retraining the fieldworkers helped improve accuracy. All statistics and plots were conducted in R version 3.4.0 [21], eCRF flowchart and EDC diagram were created using www.draw.io (JGraph, London, England).

## Results

### Data collection volume, time and accuracy

We recorded demographics of 308,348 individuals from 80,851 households in three countries between June 2016 and October 2016; 97,410 individuals and 22,364 households from Malawi, 100,207 and 32,368 from Nepal, and 110,731 and 26,119 from Bangladesh. Completeness of household demographics enumeration were 94.2%, 75.6% and 79.2% for Malawi, Nepal and Bangladesh, respectively, relative to background household count. The average number of weeks for enumeration was 14.7 range (13-16) using 20, 25, 20 field workers from Malawi, Nepal and Bangladesh, respectively. Overall, 21.7 errors (95% confidence interval: 21.4, 22.0) per 10,000 data points were found; 15.9 errors (95% confidence interval: 15.4, 16.4), 34.2 errors (95% confidence interval: 33.5, 34.9), and 14.6 errors (95% confidence interval: 14.2, 15.0) per 10,000 data points from Malawi, Nepal and Bangladesh, respectively. Of the 17,707 errors documented from all sites, the majority 12,740 (72.0%) occurred on text fields compared to numeric fields 3,868 (21.8%). In addition, 1,099 (6.2%) errors occurred as duplicate records (e.g. either by enumerating a household or any of its members at least twice) (Table 1).

**Table 1.**
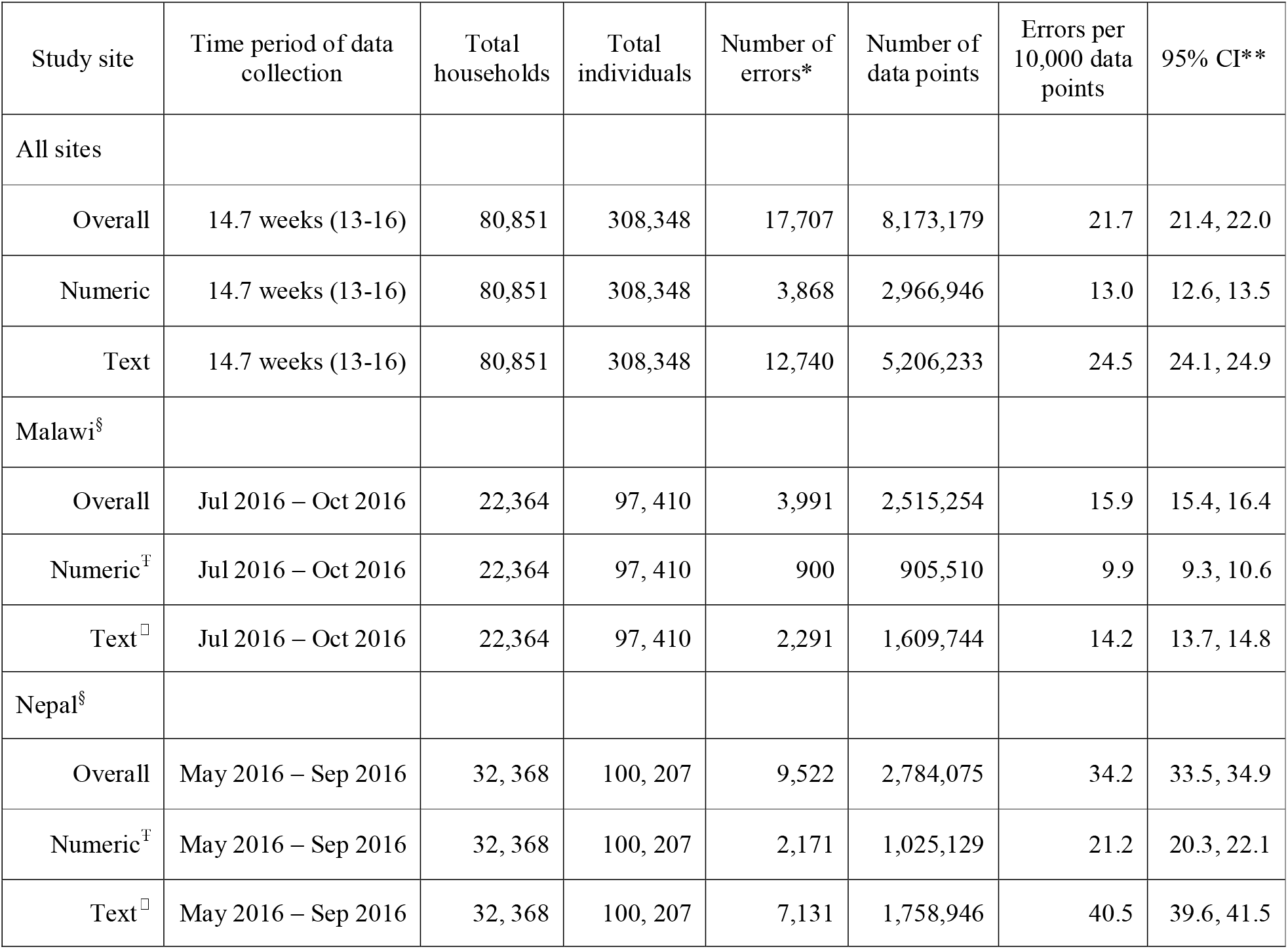
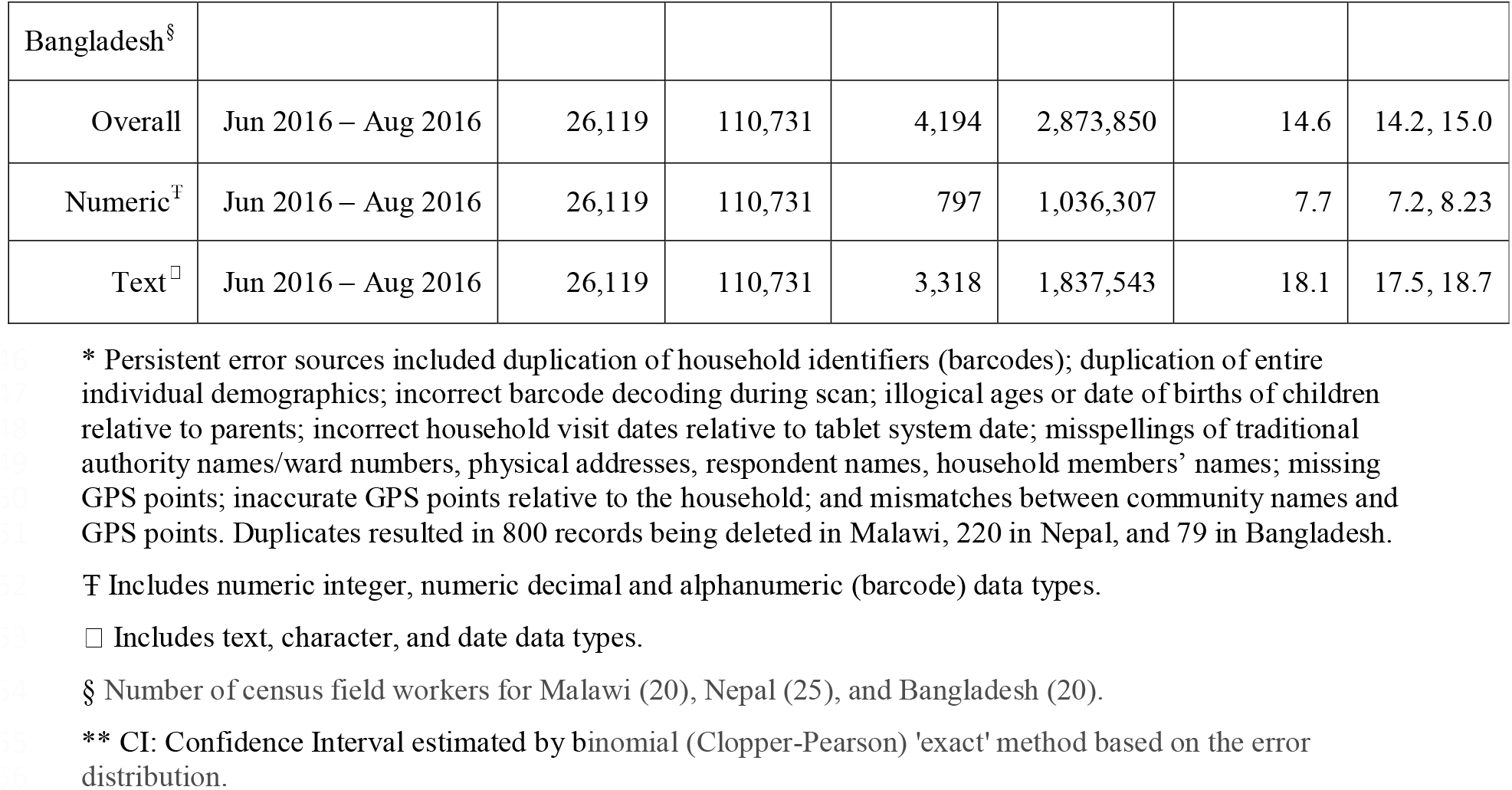
Census Data Collection Time, Volume and Accuracy in Three Typhoid Endemic Sites, 2016.

Of all the data entry errors observed during enumeration period, 2,611 (65.4%), 6,265 (65.8%) and 3,013 (71.8%) were, respectively, committed in Malawi, Nepal and Bangladesh prior fieldworkers’ retraining. Moreover, there were fewer errors observed after retraining of fieldworkers compared to pre-retraining, and the differences were statistically significant in Malawi (W =5.5, *P* <0.001), Nepal (W =19.5, *P* <0.001), and Bangladesh (W =0, *P* <0.001) (Fig. 3).

**Figure 3.**
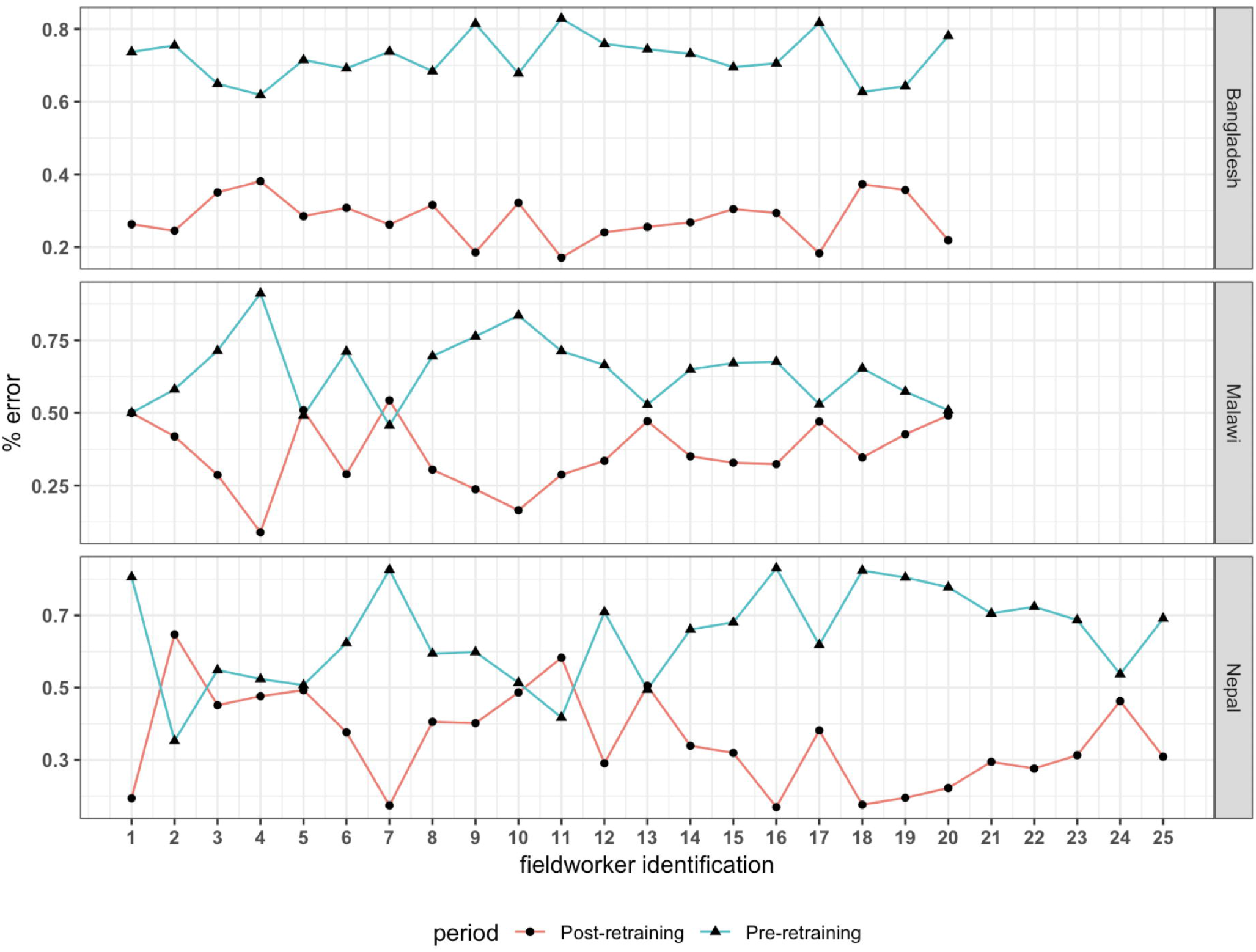
Data entry errors before and after retraining of fieldworkers, 2016.

### Time and cost of census materials

The time required to attain each material or complete each activity in preparation for census implementation varied by study site, ranging from 2 to 60 days. The most time-consuming activity was the development and customization of eCRF, which was completed in 60 days collectively. This was followed by the procurement of tablets and backpacks, which were acquired in between 7 and 60 days. In addition, we also procured and designed household identifier (barcode) stickers in between 7 and 21 days. Replacement of malfunctioned tablets reported by each study was accomplished within 30 days. We extensively trained our study fieldworkers for up to 5 days focussing on the study protocol, practical aspect of completing an eCRF, and community engagement skills. Selection of potential fieldworkers to join the study team was sorely based on successful completion of the training. Computer servers and network devices to enable data storage and transfers from tablets were pre-existing in Malawi and Bangladesh, and newly acquired in Nepal within 30 days (Table 2).

**Table 2.**
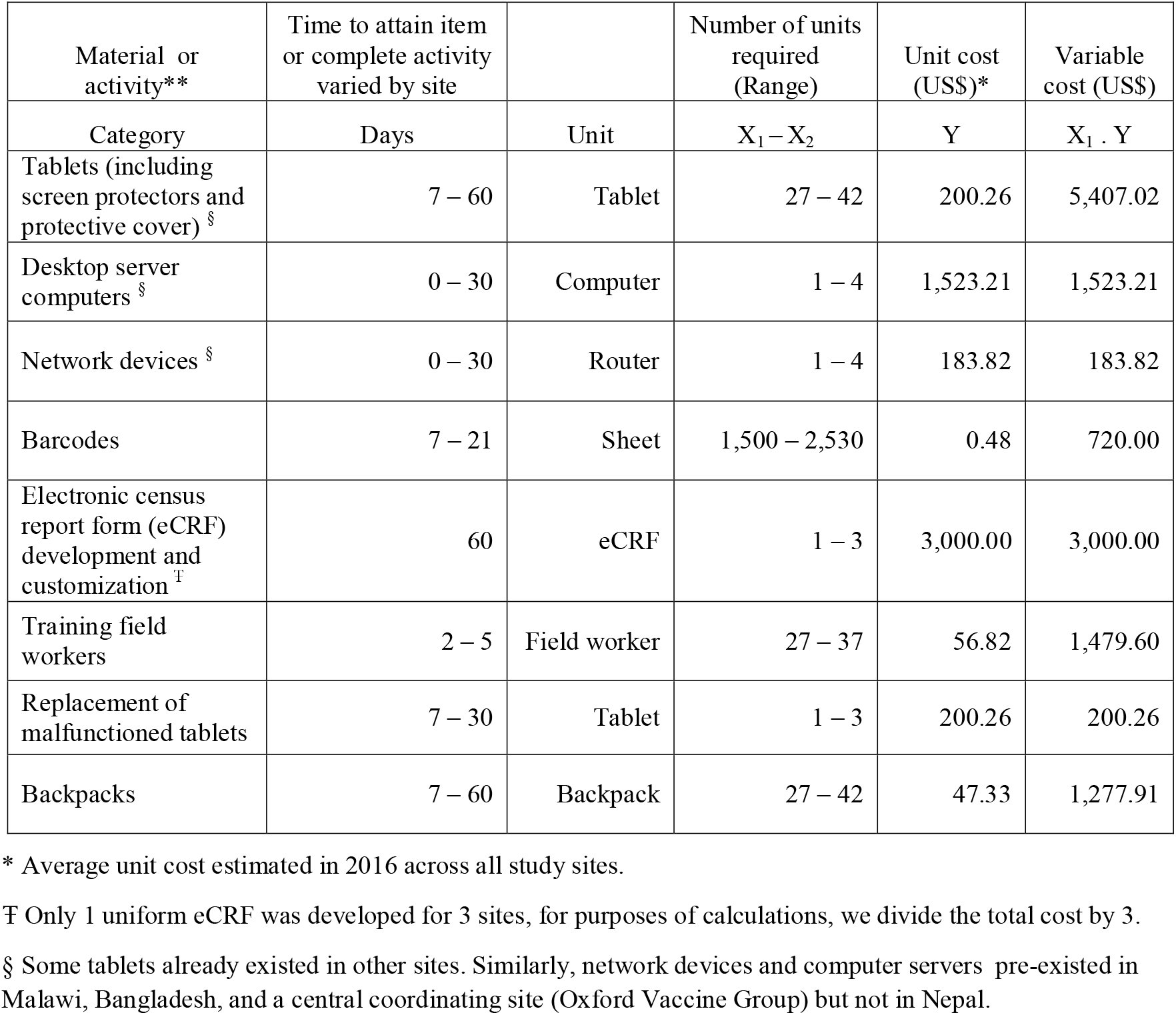
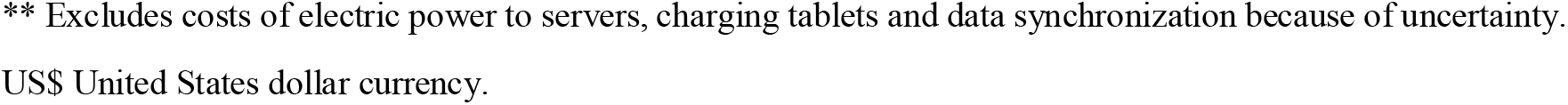
Time and Costs Attainment Prior to Implementation of an Electronic Data Capture System (EDC) in Three Typhoid Endemic Sites, 2016.

The major variable cost was incurred by customization of eCRF for use in ODK Collect for a total of US$ 9,000 for all sites, followed by procurement of 27 tablets at a variable cost of US$5,407.02. Other prominent variable costs included procurement of a desktop server (at US$1,523.21), training 27 field workers to use an eCRF and in field practices (at US$1,479.60), procurement and shipment of 27 backpacks (at US$1,277.91) and 1,500 barcode sheets (at US$720.00), replacement of a malfunctioned tablet (at US$200.26) and procurement of a network router (at $183.82). The total variable cost for the EDC was US$13,791.82 per site (Table 2).

## Discussion

In this study, we have developed and implemented an EDC which allows high volume of data collection over short time periods, high data accuracy, 12-hourly updated data access, and quality checking for decision making. Additionally, the EDC is robust, allowing for automated reports generation, scalability and could be adaptable to other epidemiological settings. Finally, the total variable cost of the EDC’s pre-census materials and activities, was minimal relative to paper-based data collection methods from similar settings.

Data were collected by largely secondary school level only fieldworkers receiving 1 week of training and a day of retraining, and although the learning curve of using an eCRF in ODK Collect on Android-based tablets was steep in the first 5 weeks of field work, high volume and fairly accurate data were recorded (Fig. 3 and Fig. 4). The data accuracy of ∼0.22% errors (21.7 errors per 10,000 data points) reported in this study meets the acceptable quality threshold of 50 errors per 10,000 data points recommended by the Society of Clinical Data Management (SCDM, McLean, Virginia, USA) [22, 23]. The highly accurate EDC data in this study is comparable to EDC data accuracies reported by the chronic disease research in South Africa (0.17%) and maternal health survey in Burkina Faso (0.24%) [24, 25]. However, our EDC data accuracy is superior to EDC data accuracies reported by the maternal health (2.8%) and neglected tropic disease surveys (5.2%) in Ethiopia, the bloodstream infections study in Zanzibar (1.0%) and the tuberculosis program in India (4.2%) [2, 3, 7, 26]. Moreover, our EDC data are more accurate in comparison to data reported from paper-based studies of maternal health (1.1%) and neglected tropical disease (6.2%) surveys in Ethiopia, bloodstream infections study in Zanzibar (7.0%), chronic disease research in South Africa (0.73%), and randomized controlled trial in Fiji (20.8%) [2–4, 7, 24]. As with previous studies [2, 22, 27], text fields of this eCRF generated more errors than numeric fields,, and suggest that such errors could be prevented in eCRF designs by minimizing the use of text fields through coding of text responses or leaving out insignificant text responses completely. The accuracy variations between EDCs are probably due to robustness of the EDC design in terms of error proofing. Robustness in the design is likely to depend on the limitations of software and hardware, and technical know-how of developers.

**Figure 4.**
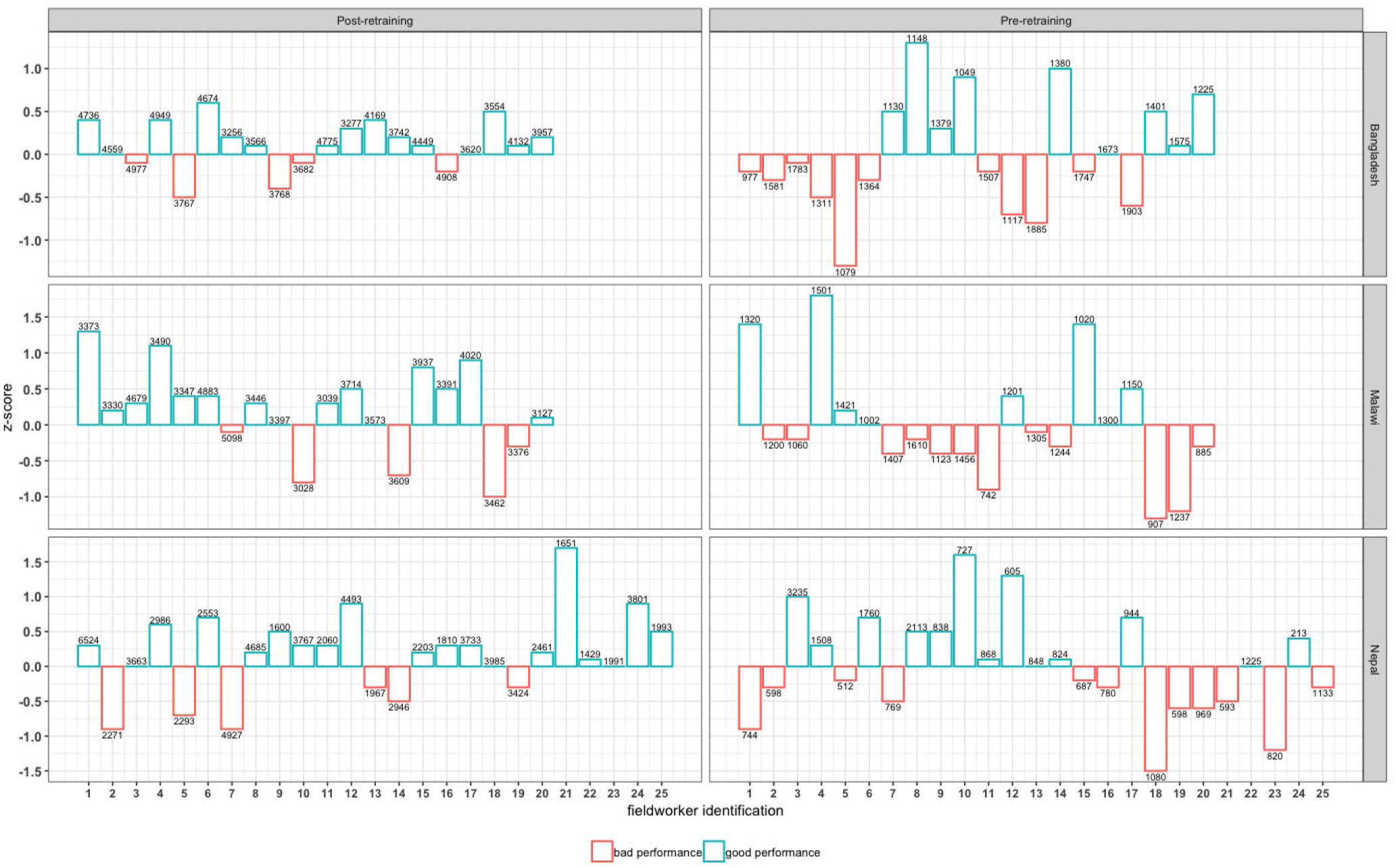
Speed and accuracy trade-off before and after retraining of fieldworkers, 2016.

Unlike the EDC and paper-based methods used in a similarly setting [28], our EDC synchronized study data updates at least every 12 hours post-data collection in order to provide recent data accessibility for decision making; Rapid accessibility to recent data has enabled immediate quality checks and data cleaning on critical variables which, at the time of the study, are beyond ODK’s built-in validations. It also enabled us to quickly understand and decide on ways to improve participant uptake rates, adding to a growing body of literature reporting how rapid data updates by an EDC enables swift decisions [9, 29, 30].

The EDC was also designed to counteract some complexities associated with data collection in low- and middle-income; Internet connectivity was through a client-server system where data capture client (ODK Collect) was an offline stand-alone instance separated from the database server (ODK Aggregate). Data were synchronized from client to server at a later point in time at the base STRATAA data office where connectivity was possible. This approach has also been recommended by others [31, 32], and we did not experience any damage or theft of the tablets which led to data loss before data was synchronized to database server. We adhered to a practice of disabling eCRF ‘edit’ options, post-interview, in order to maintain data integrity in the field. Validations within the ODK Collect prevented most errors. However, 0.4% duplicate household identities and 0.3% missing GPS points were uncovered in addition to other text and numeric errors. Following good data management practices [33], our EDC also provided three backup strategies; scheduled data synchronization to (i) centralized repository, (ii) MySQL-defined databases, and (iii) scheduled incremental backup of MySQL-defined databases to external storage devices.

The EDC delivered considerable capacity for automated report generation, scalability and adaptability. We were able to use SQL to pull seasonal data from MySQL-defined database, and automate summaries of demographics in order to monitor progress of field work, and collective and individual performance of field workers. SQL was preferred because of its simple but powerful syntax, and its wider use in handling complex queries to epidemiological datasets [22, 24, 29, 32]. Since the STRATAA consortium continuously generates laboratory data, post-census, the EDC also allows scalability, pushing laboratory data from laboratory database systems to MySQL-defined databases while keeping the database structure homogeneous across sites. The EDC could therefore not only be adopted by others collecting large data volumes requiring centralized data storage and automation of process, but also be tested by settings with little experience in conducting field-based research. The EDC is installed in three typhoid endemic settings and will be maintained by STRATAA consortium for adaptability of potential future studies.

Costs estimates on the data capture systems across low- and middle-income settings account for different item inclusions [7, 28, 34, 35]. However, generally, our total variable cost of the EDC was minimal relative to most EDCs or paper-based data collection methods conducted in similar settings. For instance, our EDC’s total variable cost is analogous to US$13,883.00 incurred on a paper-based survey of neglected tropic diseases in Ethiopia [7]. However, in northern Malawi, estimated total variable costs of an EDC (US$14,477.46 [£11,427]) and paper-based system (US$23,939.06 [£18,895]) are slightly and much higher than our EDC respectively [28]. Similarly, our total variable cost is relatively low compared to paper-based studies conducted in Bangladesh and Philippines (US$45,000.00) on verbal autopsy [34], and in Kenya (US$15,999.00) on influenza [35].

## Conclusion

In conclusion, we have designed an EDC which has been implemented in three typhoid endemic sites to collect large volume of accurate data in short time periods with rapid access through automated reports. The EDC’s development required careful attention to detail but the materials’ variable costs prior to census implementation, were minimal relative to some EDCs and paper-based data collection methods. This EDC could be adopted in similar epidemiological settings, enabling the collection and management of large data volumes, centralize data storage, and automated data processes.

## Availability and requirements

The code scripts used to develop the EDC (ODK Collect eCRF and MySQL database objects), and the raw data for errors analysed in this paper are all available through GitHub [15].

## Data Availability

The code scripts used to develop the EDC (ODK Collect eCRF and MySQL database objects), and the raw data for errors analysed in this paper are all available through GitHub.

https://github.com/Oxfordvaccinegroup/Electronic-Data-Capture-for-Large-Scale-Typhoid-Surveillance---STRATAA

## Abbreviations

EDCs: Electronic data capture systems
STRATAA: Strategic Typhoid alliance across Africa and Asia consortium
ODK: Open Data Kit
GPS: global positioning system
eCRF: electronic census report form
SQL: Structured Query Language
CI: Confidence Intervals
US$: United States dollar
SCDM: Society of Clinical Data Management

## Declarations

### Ethics approval and consent to participate

Ethical approval was obtained from the Malawi National Health Sciences Research Committee, 15/5/1599; Bangladesh ICDDR,B Institutional Review Board, PR-15119; Nepal Health Research Council, 306/2015; and Oxford Tropical Research Ethics Committee, 39-15. Following extensive sensitisation and engagement with community and traditional leaders, and community health-workers, the key informant from each household provided a verbal informed consent, to enumerate the household, which was documented in the eCRF.

### Consent for publication

Not applicable

### Availability of data and materials

The datasets generated and/or analysed during the current study are available in GitHub [15].

### Authors contribution

Conceptualization: DT, YGF, RSH; Methodology: DT, YGF, MS, NS, YA, CH; Writing-Original Draft: DT; Writing-Review and Editing: YGF, MS, NS, ST, YA, MAG, CH, JEM, AJP, RSH; Funding-Acquisition: MAG, AJP, RSH. All authors read and approved the final manuscript.

## Acknowledgements

*Members of The Strategic Typhoid alliance across Africa & Asia consortium* (*STRATAA*): Oxford University Clinical Research Unit, Patan Academy of Health Sciences, Kathmandu, Nepal (Mila Shakya, Abhilasha Karkey, Sabina Dongol, Amit Aryjal, Buddha Basnyat); International Center for Diarrhoeal Diseases Research, Dhaka, Bangladesh (Nirod Saha, Farhana Khanam, Md Arifuzzaman Khan, John D. Clemens, Firdausi Qadri, K. Zaman); Malawi Liverpool Wellcome Trust Clinical Research Programme, Malawi (Deus Thindwa, Robert S Heyderman, Melita A Gordon, Tikhala Makhaza Jere, Chisomo Msefula, Tonney Nyirenda); The Hospital for Tropical Diseases, Wellcome Trust Major Overseas Programme, Oxford University Clinical Research Unit, Ho Chi Minh City, Vietnam (Tan Trinh Van); Wellcome Trust Sanger Institute, Cambridge, United Kingdom (Stephen Reece, Gordon Dougan); Oxford Vaccine Group, Department of Paediatrics, University of Oxford, and the NIHR Oxford Biomedical Research Centre, Oxford, United Kingdom (Merryn Voysey, Christoph J. Blohmke, Jennifer Hill, Thomas C. Darton, Susan Tonks, Yama G Farooq, James E. Meiring, Andrew J Pollard); Yale School of Public Health, Yale University, New Haven, Connecticut, United States of America (Neil J. Saad, Virginia E. Pitzer); Center for Tropical Medicine and Global Health, Nuffield Department of Medicine, University of Oxford, United Kingdom (Stephen Baker, Christiane Dolecek); The Peter Doherty Institute for Infection and Immunity, The University of Melbourne, Australia (Sarah J. Dunstan); Centre for Systems Genomics, University of Melbourne, Parkville, Victoria, Australia (Kathryn E. Holt). Division of Infection and Immunity, University College London (Robert S Heyderman), Department of Infectious Disease Epidemiology, Imperial College London (Deus Thindwa), Institute of Infection and Global Health, University of Liverpool, Liverpool, United Kingdom (Melita A Gordon). We are grateful to all individuals who participated in the census of the STRATAA study. We also thank all community leaders from the STRATAA areas for allowing us to conduct STRATAA study in their areas.

